# Influenza vaccine effectiveness against pneumonia and COPD exacerbations among patients with chronic obstructive pulmonary disease in Thailand: A national test-negative design study, 2013–2024

**DOI:** 10.64898/2026.05.26.26354178

**Authors:** Sutthinan Chawalchitiporn, Pichaya Tantiyavarong, Jiraphut Kittiwatanachod, Suriya Naosri, Kriengkrai Prasert, Prabda Praphasiri

**Author notes:** Corresponding author: Prabda Praphasiri, Faculty of Public Health, Kasetsart University Chalermphrakiat Sakon Nakhon Province Campus, Sakon Nakhon, Thailand.

## Abstract

**Background/Objectives:** Influenza infection is a major trigger of pneumonia and acute exacerbations among patients with chronic obstructive pulmonary disease (COPD). However, national laboratory-confirmed evidence on influenza vaccine effectiveness (VE) in this high-risk population remains limited. This study aimed to estimate the effectiveness of seasonal influenza vaccination against influenza-associated pneumonia and COPD exacerbations among patients with COPD in Thailand.

**Methods:** We conducted a nationwide retrospective test-negative design study using administrative healthcare data from the National Health Security Office linked with laboratory-confirmed influenza surveillance data between June 1, 2013, and May 31, 2025, covering twelve influenza seasons (2013–2024). COPD-related clinical episodes among patients aged ≥40 years who presented with pneumonia or acute exacerbation of COPD and underwent RT-PCR testing for influenza were included. Multilevel Poisson regression models were used to estimate adjusted risk ratios (RRs), and VE was calculated as (1 − adjusted RR) × 100.

**Results:** A total of 606,072 COPD-related clinical episodes were included, of which 192,224 (31.7%) were influenza-positive. The overall adjusted VE against influenza-associated pneumonia was 63.2% (95% CI: 62.5–64.0), while VE against influenza-associated COPD exacerbations was 67.0% (95% CI: 48.8–78.8). VE estimates were broadly similar across age groups and remained substantial across COPD severity strata. Although point estimates were numerically higher in severe and very severe COPD, subgroup differences should be interpreted cautiously.

**Conclusions:** Seasonal influenza vaccination was associated with substantial protection against influenza-associated pneumonia and COPD exacerbations among patients with COPD in Thailand.

## INTRODUCTION

Chronic obstructive pulmonary disease (COPD) is a major cause of morbidity and mortality worldwide and represents a substantial public health burden [1–3]. Globally, COPD ranks among the leading causes of death and is projected to increase further due to population ageing and continued exposure to risk factors such as smoking and air pollution [2,3]. In Thailand, COPD contributes significantly to hospital admissions and long-term disability, particularly among older adults, placing considerable pressure on healthcare systems [4,5].

Influenza infection is a well-recognized trigger of acute respiratory deterioration in patients with COPD. Viral infection can exacerbate airway inflammation, impair mucociliary clearance, and increase susceptibility to secondary bacterial infection, thereby precipitating acute exacerbations and pneumonia [6–9]. These complications are associated with increased hospitalization, healthcare utilization, and excess mortality [10,11]. Preventing influenza infection among individuals with COPD is therefore an important strategy to reduce respiratory morbidity and mortality.

Annual influenza vaccination is recommended for individuals with chronic respiratory diseases by the World Health Organization and multiple international clinical guidelines [12–14]. In Thailand, influenza vaccination has been incorporated into the national immunization program for high-risk populations, including patients with COPD, under the Universal Coverage Scheme [15]. Despite these policy efforts, vaccination coverage among individuals with chronic diseases has historically remained suboptimal [16]. Generating robust population-level evidence on vaccine effectiveness (VE) is therefore essential to inform public health decision-making and strengthen vaccination strategies.

Several studies have evaluated influenza vaccine effectiveness among patients with COPD [9,17]. Although many have reported protective effects against respiratory outcomes, a substantial proportion relied on clinically diagnosed outcomes without laboratory confirmation, which may introduce outcome misclassification. In addition, retrospective cohort studies based solely on administrative data may be limited when complete person-time information is unavailable or when healthcare-seeking behavior differs between vaccinated and unvaccinated individuals.

The test-negative design (TND) has emerged as a widely used approach for evaluating influenza VE in real-world settings [18,19]. By comparing vaccination status between patients who test positive and those who test negative for influenza among individuals presenting with similar respiratory syndromes, the TND helps reduce bias related to healthcare-seeking behavior and testing practices. When combined with laboratory confirmation using reverse transcription polymerase chain reaction (RT-PCR), this design can provide more valid and reliable estimates of VE against influenza-associated outcomes.

However, national laboratory-confirmed evidence on influenza VE among patients with COPD in Thailand remains limited. In addition, few studies have examined VE separately for influenza-associated pneumonia and COPD exacerbations or explored potential variation by age, disease severity, influenza activity level, and viral strain.

Therefore, this study aimed to estimate the effectiveness of seasonal influenza vaccination against laboratory-confirmed influenza-associated pneumonia and COPD exacerbations among patients with COPD in Thailand between 2013 and 2024 using nationwide linked administrative and surveillance data. By providing national-level evidence across multiple influenza seasons, the findings may help inform public health policy and optimize influenza prevention strategies for high-risk populations in Thailand.

## MATERIALS AND METHODS

### Study design and data sources

We conducted a nationwide retrospective study using a test-negative design (TND) to estimate influenza vaccine effectiveness among patients with chronic obstructive pulmonary disease (COPD) in Thailand. Administrative healthcare records from the National Health Security Office (NHSO) were linked with laboratory-confirmed influenza surveillance data from the Department of Medical Sciences (DMS) between June 1, 2013, and May 31, 2025, covering twelve influenza seasons. Seasons were labeled by the calendar year in which they began; accordingly, the final (2024) season ran from June 1, 2024 to May 31, 2025.

Data were accessed for research purposes in May 2026. Authors did not have access to information that could identify individual participants during or after data collection, as all records were de-identified prior to linkage.

The NHSO database contains electronic medical records from public hospitals under Thailand’s Universal Coverage Scheme and includes demographic characteristics, diagnostic codes based on the International Classification of Diseases, 10th Revision (ICD-10), healthcare utilization data, and vaccination records. Influenza infection was confirmed using reverse transcription polymerase chain reaction (RT-PCR) testing performed by the DMS. Influenza activity data were obtained from national influenza surveillance records from the Department of Medical Sciences. Influenza seasons were defined from June 1 to May 31 of the following year, and each season was labeled according to the starting calendar year (e.g., the 2013 season corresponds to June 1, 2013–May 31, 2014).

### Study population

Eligible episodes were identified among patients aged ≥40 years with a diagnosis of COPD (ICD-10 codes J40, J44.0, J44.1) who presented with either: 1) pneumonia (ICD-10 J12–J18), or 2) acute exacerbation of COPD (ICD-10 J44.0 or J44.1), and underwent RT-PCR testing for influenza during the same clinical episode. Eligibility criteria for the analytic dataset required age ≥40 years, absence of malignancy or immunocompromising conditions, and complete key covariates required for severity classification. The age threshold of 40 years was applied to improve the specificity of COPD case identification in administrative data, because COPD diagnosed before age 40 years is uncommon and may reflect alternative chronic airway disorders. Episodes were classified as:

- **Cases:** RT-PCR positive for influenza (A or B);
- **Controls:** RT-PCR negative for influenza.

The unit of analysis was the COPD-related clinical episode, and all analyses were therefore conducted at the episode level rather than the individual-patient level. The TND approach assumes that cases and controls seek care for similar respiratory syndromes and have comparable access to testing, thereby minimizing healthcare-seeking bias.

### Exposure definition

Receipt of seasonal influenza vaccination was identified from NHSO vaccination records. Vaccinated individuals were defined as those with a recorded seasonal influenza vaccination during the same influenza season and at least 14 days before the date of specimen collection for influenza testing. Individuals without a recorded vaccination during the same season, or vaccinated less than 14 days before testing, were classified as unvaccinated.

### Outcomes

Two primary outcomes were evaluated separately:

- Laboratory-confirmed influenza-associated pneumonia;
- Laboratory-confirmed influenza-associated COPD exacerbations.

Outcome status was determined based on RT-PCR positivity among patients presenting with the respective clinical diagnoses.

### Covariates

Potential confounders were selected based on clinical relevance and prior literature. The following variables were included as fixed effects in the multivariable models: 1) age group (40–59, 60–74, ≥75 years); 2) sex; 3) comorbidity status; 4) smoking status (never, former, current); 5) COPD severity level; 6) healthcare utilization in the prior year; 7) influenza activity level; and 8) influenza season (2013–2024). Comorbidity status was classified as the presence or absence of major comorbid conditions, including cardiovascular disease, diabetes mellitus, hypertension, or other chronic diseases. Smoking status was categorized as never, former, or current. Healthcare utilization was defined as the total number of outpatient and inpatient visits during the preceding 12 months and was included as a proxy measure of underlying healthcare contact and access. For analysis, this variable was grouped into low, medium, and high utilization strata. COPD severity was classified using an adapted framework informed by GOLD staging principles, applied to available administrative data elements including prescribed inhaled therapy class, recorded mMRC dyspnea and CAT scores where present, prior exacerbation history, and documentation of respiratory failure or heart failure. Because spirometric data are not captured in NHSO administrative records, this classification represents a proxy measure and does not constitute validated GOLD spirometric staging; misclassification of severity category cannot be excluded, although the ordinal ranking is expected to reflect meaningful clinical gradients in disease burden. Influenza activity level was defined using national surveillance data from the Department of Medical Sciences according to the World Health Organization threshold method. Under this framework, influenza activity is classified into escalating epidemic thresholds. Because no analytic episodes occurred during alert-threshold periods in the present dataset, activity level was categorized as low, moderate, or high in the analysis.

### Statistical analysis

Vaccine effectiveness (VE) was estimated using multilevel generalized linear models with a Poisson distribution and log link to directly estimate adjusted risk ratios (RRs). Province was included as a random intercept to account for clustering within geographic areas. Robust standard errors were applied to account for potential variance misspecification using the modified Poisson approach described by Zou [20].

Risk ratios were estimated instead of odds ratios because laboratory-confirmed influenza was not a rare outcome in this population. In such settings, odds ratios may overestimate the magnitude of association. Direct estimation of RR provides a more interpretable measure of vaccine effectiveness, calculated as:

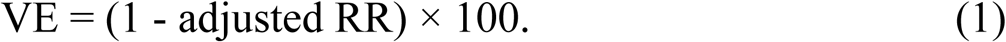

Primary analyses were conducted separately for pneumonia and COPD exacerbations. Secondary analyses included subgroup analyses by age group, COPD severity, influenza activity level, and viral strain. Year-specific VE estimates were derived using interaction terms between vaccination status and influenza season. Statistical significance was defined as a two-sided p-value < 0.05. In subgroup analyses where multilevel models did not converge, Poisson regression models with cluster-robust standard errors at the province level were used as an alternative approach.

All analyses were performed using Stata version 19.5 SE (StataCorp LLC, College Station, TX, USA).

### Ethical considerations

The study was conducted in accordance with the Declaration of Helsinki and approved by the Research Ethics Committee of Thammasat University Faculty of Medicine (Approval No. 138/2569). Patient consent was waived due to the use of de-identified administrative data.

## RESULTS

### Study population

Between June 1, 2013, and May 31, 2025, a total of 606,072 COPD-related clinical episodes that underwent RT-PCR testing for influenza were included in the test-negative design analysis (Fig 1). Of these, 413,848 episodes (68.3%) were influenza-negative, and 192,224 episodes (31.7%) were influenza-positive. The relatively high overall positivity proportion reflects the study design: the analytic dataset was restricted to RT-PCR-tested episodes, and testing in this setting was concentrated among patients presenting with acute respiratory syndromes during periods of active influenza surveillance, enriching the tested population for influenza-associated illness relative to an unselected clinical population.

**Fig 1.**
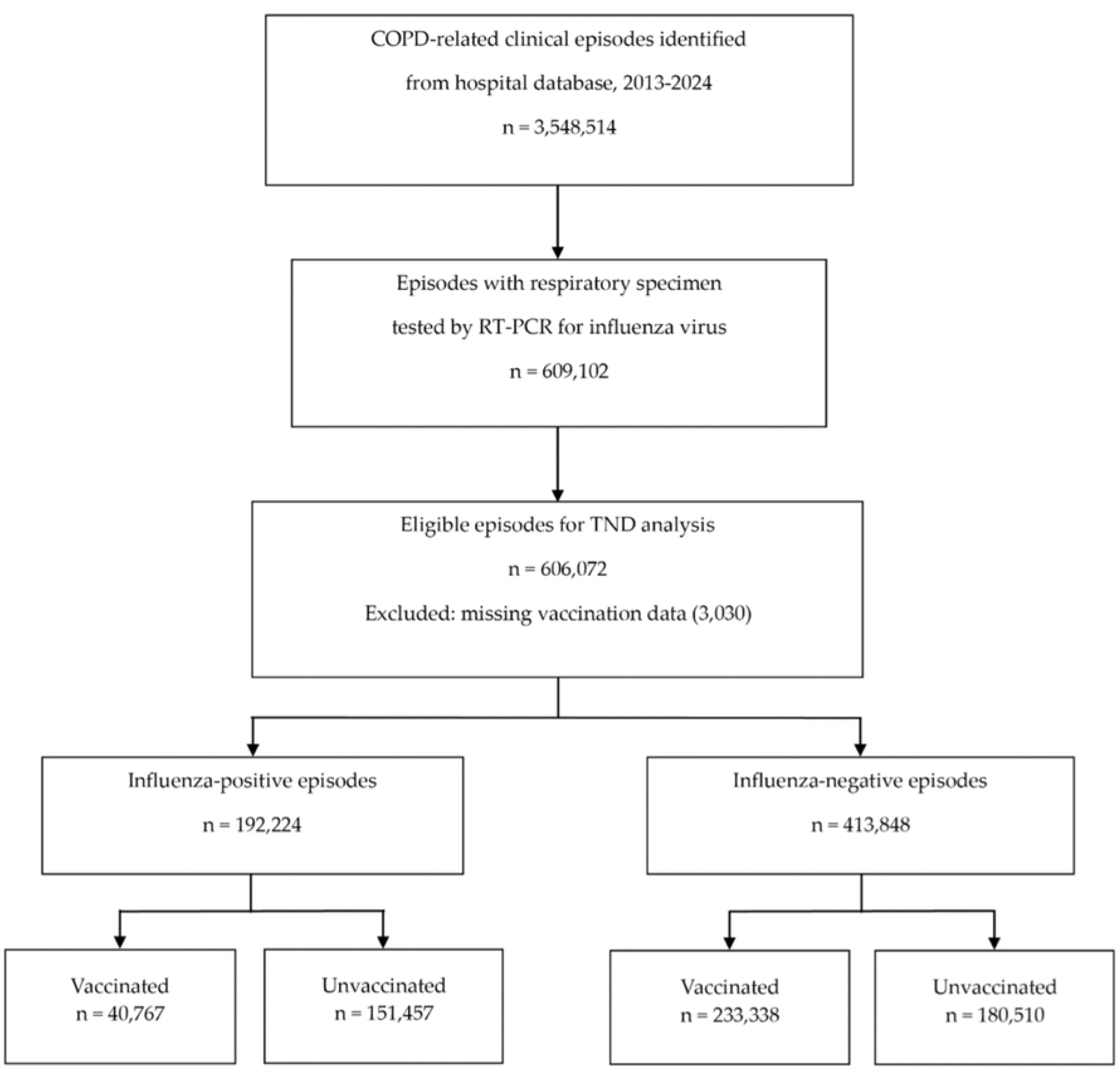
Study flow diagram of COPD-related clinical episodes included in the test-negative design analysis, Thailand, 2013–2024. Percentages are calculated within each relevant denominator.

The mean age differed slightly between influenza-negative and influenza-positive episodes (69.4 ± 10.8 vs. 69.1 ± 11.3 years; p < 0.001), although the effect size was small (Cohen’s d = 0.031; 95% CI: 0.026–0.035). The distribution of age groups also differed between groups (p < 0.001), with a slightly higher proportion of influenza-positive episodes occurring among patients aged 40–59 years (34.2% vs. 33.1%; standardized difference = 0.03).

Sex distribution was similar between groups, although the overall difference was statistically significant (p < 0.001; standardized difference = 0.02), with males accounting for most episodes (78.5% vs. 78.1%). Male episodes predominated in both influenza-negative and influenza-positive groups, suggesting that the study sample was heavily male rather than that influenza positivity differed materially by sex.

Influenza-positive episodes were more likely to occur among patients with comorbid conditions (8.8% vs. 7.8%; standardized difference = 0.05) and among current smokers (8.4% vs. 7.5%; standardized difference = 0.04). Regarding disease severity, influenza-positive episodes occurred slightly more frequently among patients with severe or very severe COPD (24.1% vs. 23.2%; standardized difference = 0.03). Healthcare utilization in the prior year also differed modestly between groups (standardized difference = 0.06). Vaccination during the same influenza season was substantially lower among influenza-positive episodes than among influenza-negative episodes (21.2% vs. 56.4%; standardized difference = 0.49; p < 0.001). The distribution of influenza activity levels differed modestly between groups (p = 0.027; standardized difference = 0.02) (Table 1).

**Table 1.** Baseline characteristics of RT-PCR tested COPD episodes by influenza test result, Thailand, 2013–2024.

| Characteristic | Influenza-negative<br>(n=413,848) | Influenza-positive<br>(n=192,224) | p-value | Effect size |
| --- | --- | --- | --- | --- |
| **Age, mean (SD)** | 69.4 (10.8) | 69.1 (11.3) | <0.001 | 0.031<br>(0.026–0.035) |
| **Age group, n (%)** |  |  |  |  |
| 40–59 years | 136,798 (33.1) | 65,785 (34.2) |  |  |
| 60–74 years | 130,453 (31.5) | 57,697 (30.0) | <0.001 | 0.03 |
| ≥75 years | 146,597 (35.4) | 68,742 (35.8) |  |  |
| **Sex, n (%)** |  |  |  |  |
| Female | 90,611 (21.9) | 41,405 (21.5) |  |  |
| Male | 323,237 (78.1) | 150,819 (78.5) | <0.001 | 0.02 |
| **Comorbidity status, n (%)** |  |  |  |  |
| No | 381,685 (92.2) | 175,330 (91.2) |  |  |
| Yes | 32,163 (7.8) | 16,894 (8.8) | <0.001 | 0.05 |
| **Smoking status, n (%)** |  |  |  |  |
| Never | 89,089 (21.5) | 42,627 (22.2) |  |  |
| Former | 293,761 (71.0) | 133,401 (69.4) | <0.001 | 0.04 |
| Current | 30,998 (7.5) | 16,196 (8.4) |  |  |
| **COPD severity, n (%)** |  |  |  |  |
| Mild | 225,431 (54.5) | 103,692 (53.9) |  |  |
| Moderate | 92,279 (22.3) | 42,358 (22.0) |  |  |
| Severe | 57,617 (13.9) | 27,981 (14.6) | <0.001 | 0.03 |
| Very severe | 38,521 (9.3) | 18,193 (9.5) |  |  |
| **Healthcare utilization (prior year), n (%)** |  |  |  |  |
| Low | 120,477 (29.1) | 60,508 (31.5) |  |  |
| Medium | 128,676 (31.1) | 60,649 (31.5) | <0.001 | 0.06 |
| High | 164,695 (39.8) | 71,067 (37.0) |  |  |
| Vaccinated in the same season, n (%) | 233,338 (56.4) | 40,767 (21.2) | <0.001 | 0.49 |
| Influenza activity level, n (%) |  |  |  |  |
| Low | 165,342 (40.0) | 70,383 (36.6) |  |  |
| Moderate | 238,416 (57.6) | 116,992 (60.9) | 0.027 | 0.02 |
| High | 10,090 (2.4) | 4,849 (2.5) |  |  |

**Abbreviations:** CI, Confidence Interval; COPD, Chronic Obstructive Pulmonary Disease; GOLD, Global Initiative for Chronic Obstructive Lung Disease; SD, Standard Deviation.
**Notes:** Values are presented as mean (SD) or n (%). P-values were calculated using a two-sample t-test for age and chi-square tests for categorical variables. Effect size for age is reported as Cohen's d with 95% CI; effect sizes for categorical variables are reported as standardized differences. COPD severity was classified according to an adapted GOLD-based framework using available information on prescribed inhaled therapy, mMRC dyspnea score, CAT score, prior exacerbation history, and markers of advanced disease such as respiratory failure or heart failure.

### Overall vaccine effectiveness

In multilevel Poisson regression models adjusted for demographic and clinical covariates, seasonal influenza vaccination was significantly associated with reduced risk of both influenza-associated pneumonia and COPD exacerbations.

Across twelve influenza seasons (2013–2024), the overall adjusted vaccine effectiveness (VE) was 63.2% (95% CI: 62.5–64.0) against influenza-associated pneumonia and 67.0% (95% CI: 48.8–78.8) against influenza-associated COPD exacerbations (Table 2).

**Table 2.** Vaccine effectiveness overall and by subgroup among patients with COPD, Thailand, 2013–2024.

| Category | Pneumonia<br>VE % (95% CI) | COPD exacerbations<br>VE % (95% CI) |
| --- | --- | --- |
| Overall | 63.2 (62.5–64.0) | 67.0 (48.8–78.8) |
| Age group |  |  |
| 40–59 years | 62.8 (61.5–64.3) | 66.1 (64.8–67.5) |
| 60–74 years | 65.5 (64.0–67.0) | 65.5 (64.0–66.9) |
| ≥75 years | 63.5 (62.0–65.0) | 65.6 (64.3–66.9) |
| **COPD severity** |  |  |
| Mild | 61.1 (59.9–62.1) | 66.7 (48.5–78.4) |
| Moderate | 60.3 (58.6–61.9) | 63.4 (61.6–65.0) |
| Severe | 70.5 (68.8–72.1) | 76.4 (75.0–78.0) |
| Very severe | 69.3 (67.1–71.3) | 71.6 (70.0–74.0) |

**Abbreviations:** CI, Confidence Interval; COPD, Chronic Obstructive Pulmonary Disease; VE, Vaccine Effectiveness.
**Notes:** VE was calculated as $(1 - \text{adjusted risk ratio}) \times 100$ . Adjusted risk ratios were estimated using multilevel Poisson regression models with province included as a random intercept. Overall models were adjusted for age, sex, comorbidity status, smoking status, COPD severity, healthcare utilization, and influenza activity level. Stratified models were adjusted for the same covariates except the stratification variable.

### Vaccine effectiveness by age group

VE estimates were broadly similar across age strata. For influenza-associated pneumonia, VE ranged from 62.8% (95% CI: 61.5–64.3) among patients aged 40–59 years to 65.5% (95% CI: 64.0–67.0) among those aged 60–74 years and was 63.5% (95% CI: 62.0–65.0) among those aged ≥75 years. For COPD exacerbations, VE ranged from 65.5% to 66.1% across age groups (Table 2).

### Vaccine effectiveness by COPD severity

Vaccine effectiveness remained substantial across all COPD severity strata. For pneumonia, VE ranged from 60.3% (95% CI: 58.6–61.9) in patients with moderate disease to 70.5% (95% CI: 68.8–72.1) in those with severe disease and 69.3% (95% CI: 67.1–71.3) in those with very severe COPD. For COPD exacerbations, VE ranged from 63.4% (95% CI: 61.6–65.0) among patients with moderate disease to 76.4% (95% CI: 75.0–78.0) among those with severe disease, while VE among patients with very severe COPD was 71.6% (95% CI: 70.0–74.0). Although point estimates were numerically higher in severe and very severe COPD, these subgroup differences should be interpreted cautiously and should not be taken to indicate a clear biologic gradient of increasing vaccine benefit with increasing disease severity (Table 2).

### Year-specific vaccine effectiveness

Year-specific analyses demonstrated variation in vaccine effectiveness across influenza seasons (Fig 2). For pneumonia, VE ranged from 50.2% in 2021 to 78.6% in 2013. For COPD exacerbations, VE ranged from 50.2% in 2014 to 73.8% in 2013. Protective effects of vaccination were observed in all influenza seasons evaluated.

**Fig 2.**
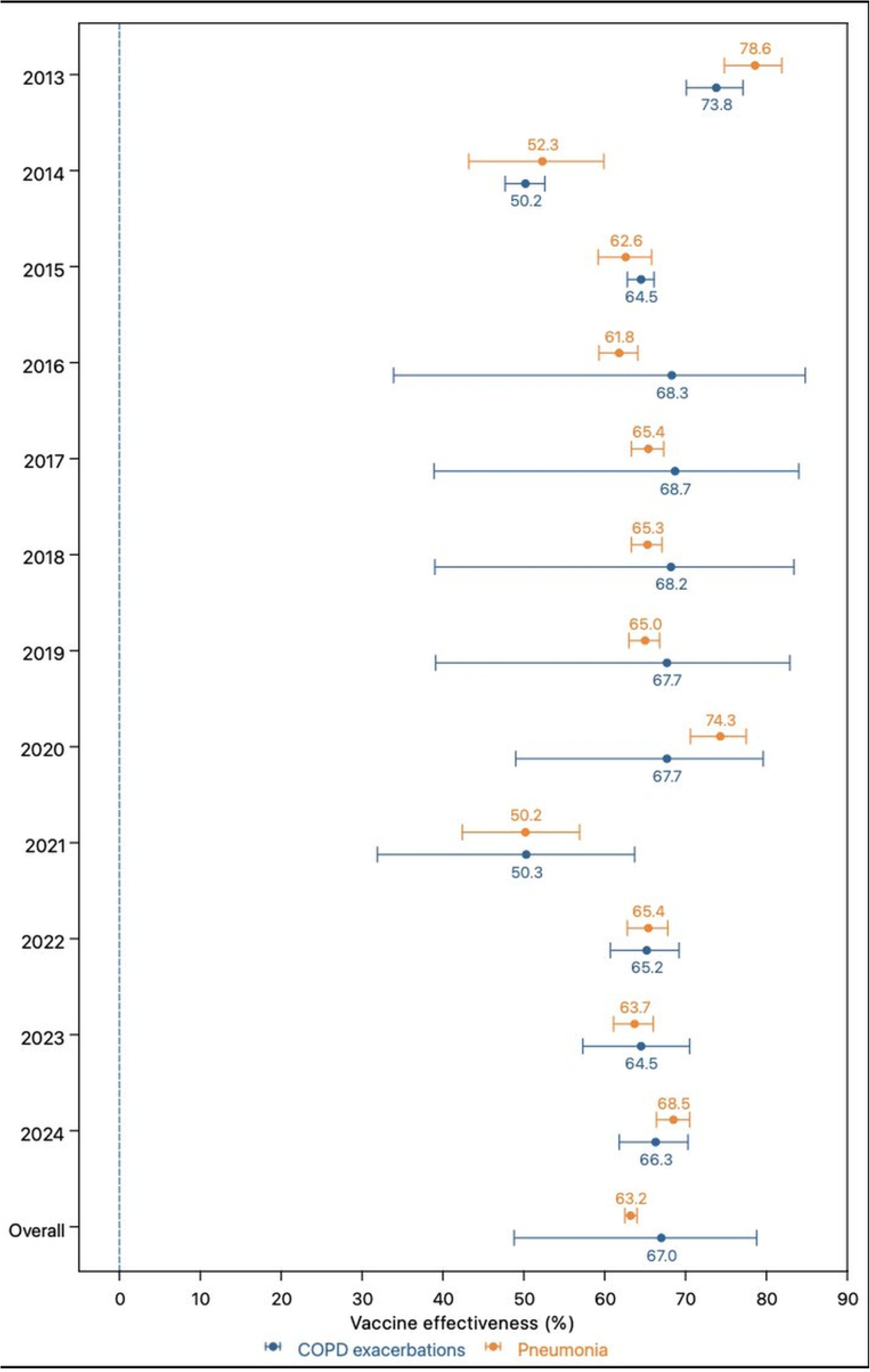
Year-specific vaccine effectiveness against influenza-associated pneumonia and COPD exacerbation among patients with COPD, Thailand, 2013–2024. Points represent vaccine effectiveness estimates, and horizontal bars indicate 95% confidence intervals.

### Vaccine effectiveness by influenza activity level

When stratified by influenza activity level, VE against pneumonia remained relatively consistent across activity levels, ranging from 61.5% during high activity periods to 63.5% during moderate activity periods. For COPD exacerbations, VE was 73.8% (95% CI: 70.1–77.1) during low activity periods and 66.3% (95% CI: 61.8–70.3) during high activity periods; the estimate during moderate activity periods (67.7%; 95% CI: 39.1–82.9) was imprecise and should be interpreted cautiously (S1 Table).

### Strain-specific vaccine effectiveness

Strain-specific analyses demonstrated variation in vaccine effectiveness (S2 Table). For pneumonia, VE was highest against influenza B (85.0%; 95% CI: 84.1–85.8), followed by A(H1N1) (65.7%; 95% CI: 64.2–67.2) and A(H3N2) (51.6%; 95% CI: 50.4–52.8). For COPD exacerbations, VE was also highest against influenza B (84.2%; 95% CI: 79.3–87.9). VE estimates against A(H1N1) (61.8%; 95% CI: 29.7–79.3) and A(H3N2) (58.9%; 95% CI: 29.6–76.0) were directionally consistent but had wide confidence intervals reflecting limited precision in seasons with lower subtype circulation and fewer COPD exacerbation episodes per strain stratum; these estimates should be interpreted cautiously.

## DISCUSSION

In this nationwide test-negative design study covering twelve influenza seasons (2013–2024), seasonal influenza vaccination was associated with substantial protection against both influenza-associated pneumonia and COPD exacerbations among patients with chronic obstructive pulmonary disease (COPD) in Thailand. The overall adjusted vaccine effectiveness (VE) was 63.2% against pneumonia and 67.0% against COPD exacerbations. These findings provide national laboratory-confirmed evidence supporting influenza vaccination in a high-risk population using linked administrative healthcare and surveillance data.

The magnitude of vaccine effectiveness observed in this study is consistent with previous research demonstrating protective effects of influenza vaccination among patients with COPD [9,17]. Notably, the meta-analysis by Bao et al. [9] reported that influenza vaccination reduced the risk of COPD exacerbations and hospitalizations, and specifically identified greater benefit among patients with severe airflow limitation (GOLD stages 3–4) — a finding directionally consistent with the numerically higher VE point estimates observed in the severe and very severe COPD subgroups in the present study, though subgroup differences should be interpreted with caution given heterogeneity in study design, outcome definitions, and analytic approaches across the literature [9,17–19]. Compared with most studies included in that meta-analysis, which relied on clinically defined outcomes without laboratory confirmation, the present study used RT-PCR–confirmed influenza as the outcome criterion, reducing outcome misclassification and providing a more specific estimate of VE against influenza-attributable disease. Influenza infection is a well-established trigger of acute COPD exacerbations and lower respiratory tract complications. Viral infection can amplify airway inflammation, impair mucociliary clearance, and increase susceptibility to secondary bacterial infection. By preventing influenza infection, vaccination may reduce viral-induced respiratory deterioration and subsequent clinical complications among patients with COPD [6–9].

Our findings are broadly consistent with prior evidence showing that influenza vaccination reduces adverse respiratory outcomes among patients with COPD [9,17]. Compared with prior Thai retrospective cohort evidence [17], the present study adds national laboratory-confirmed evidence using a test-negative design, which helps reduce bias related to healthcare-seeking behavior and testing practices [18,19]. Direct quantitative comparison should nevertheless be made cautiously because previous studies have differed in outcome definitions, study populations, healthcare settings, and analytic methods [9,17–19].

In our study, vaccination was associated with protection against both pneumonia and COPD exacerbations, with slightly higher effectiveness observed for pneumonia prevention. Differences between outcomes may reflect variation in underlying pathophysiological mechanisms. Acute exacerbations of COPD may be triggered by multiple infectious and non-infectious factors, whereas influenza-associated pneumonia may be more directly related to viral infection. Therefore, the observed differences in VE between outcomes are biologically plausible.

The wider confidence intervals observed for vaccine effectiveness against COPD exacerbations, particularly in some year-specific estimates shown in Fig 2, likely reflect a combination of lower effective event counts within stratified analyses, greater clinical heterogeneity of the outcome, and potential non-differential misclassification. Unlike influenza-associated pneumonia, COPD exacerbations may be triggered by multiple influenza and non-influenza factors, including environmental exposures and other respiratory infections, which may dilute the measurable vaccine effect and increase uncertainty. In addition, because the models accounted for clustering at the provincial level, the wider confidence intervals for COPD exacerbations may also reflect underlying between-province heterogeneity.

Year-specific analyses demonstrated variability in vaccine effectiveness across influenza seasons. Such variability has been widely reported in influenza vaccine effectiveness studies and may be influenced by several factors, including vaccine-strain correspondence, circulating subtype predominance, antigenic drift, and population immunity [21]. Despite seasonal fluctuations, vaccination was associated with protective effects in all influenza seasons evaluated.

To provide descriptive context for the observed year-to-year variation in vaccine effectiveness, we summarized the available national information on vaccine components and circulating influenza strain patterns for the years in which these data could be assembled in a consistent analyzable format (S3 Table and S4 Table). These summaries showed variation in predominant circulating strains and in their correspondence with vaccine strains across 2011–2015, which may partly explain variation in vaccine effectiveness. For the present 2013–2024 study, only the overlapping study seasons were used to inform interpretation. However, because comparable strain-matching information was not available in a standardized format for all study seasons from 2013 to 2024, these data should be interpreted as contextual support rather than as a formal year-by-year strain-match analysis across the full study period. Among the three overlapping seasons for which strain composition data were available (2013–2015), a directional pattern consistent with the known influence of vaccine-strain correspondence on VE was observed: the 2013 season, characterized by predominant A(H3N2) circulation with good vaccine-component correspondence, coincided with the highest year-specific VE estimates for both outcomes in Fig 2; the 2014 season, in which circulating A(H3N2) strains showed partial divergence from the vaccine component, corresponded to markedly attenuated VE estimates. Although this observation spans only three overlapping seasons and should not be overinterpreted, it is directionally consistent with vaccine-strain correspondence as a contributing factor to year-specific VE variation in this study.

A sensitivity analysis excluding the 2020–2022 seasons yielded overall VE estimates consistent with the primary analysis, suggesting that pandemic-era disruptions to influenza surveillance and healthcare-seeking did not materially bias the pooled findings. A decline in the number of eligible episodes was observed during the 2019–2022 period, which coincided with the COVID-19 pandemic. Non-pharmaceutical interventions implemented to control SARS-CoV-2 transmission, including mask use, social distancing, and reduced population mobility, substantially reduced influenza circulation in many countries [22]. These changes may also have influenced healthcare-seeking behavior and influenza testing patterns, potentially contributing to the smaller number of episodes and greater variability in VE estimates during these seasons.

Strain-specific analyses demonstrated heterogeneity in vaccine effectiveness. Protection was highest against influenza B. For pneumonia, VE was higher against A(H1N1) than A(H3N2), whereas for COPD exacerbations the estimates for A(H1N1) and A(H3N2) were broadly similar. Differences in subtype-specific VE have been reported in previous studies and may reflect antigenic evolution of circulating strains as well as variability in vaccine strain match across seasons [21]. Continued monitoring of subtype-specific vaccine effectiveness remains important for informing vaccine strain selection and vaccination policy. Importantly, these subtype-specific differences should not be interpreted as reflecting subtype biology alone, because observed vaccine effectiveness is also influenced by the degree of correspondence between vaccine strains and the strains circulating in a given season [21].

The comparatively stable pattern of vaccine effectiveness against COPD exacerbations across seasons should also be interpreted in light of the multifactorial nature of exacerbations. Unlike laboratory-confirmed influenza-associated pneumonia, COPD exacerbations may be precipitated by multiple viral and bacterial pathogens, environmental exposures, and non-infectious triggers, so the measured vaccine effect may reflect both prevention of influenza infection itself and attenuation of downstream influenza-associated respiratory destabilization [6–9,11,21]. This may contribute to a pattern in which vaccine effectiveness remains directionally protective across seasons even when the apparent degree of vaccine-strain correspondence varies.

VE estimates remained substantial across age groups and COPD severity strata. Although some subgroup point estimates were higher in severe and very severe COPD, these findings should be interpreted cautiously. Differences across severity strata may reflect variation in case mix, healthcare utilization, residual confounding, or the precision of subgroup estimates rather than a true biologic gradient of greater vaccine responsiveness with increasing disease severity. From a clinical and public health perspective, the key finding is that meaningful protection was observed across the age and COPD severity strata represented in this study. These findings support current recommendations prioritizing influenza vaccination for adults with chronic respiratory diseases across the age strata represented in this study and across COPD severity levels [12–14].

When stratified by influenza activity level, vaccine effectiveness against pneumonia remained relatively consistent across activity periods, whereas VE against COPD exacerbations was highest during low activity periods and modestly lower during moderate and high activity periods. Differences in VE across activity levels may reflect variation in viral circulation intensity, case mix during peak influenza transmission, or potential co-infections.

This study has several strengths. First, it utilized a large national dataset linking administrative healthcare records with laboratory-confirmed RT-PCR influenza surveillance data, reducing outcome misclassification. Second, the test-negative design minimized bias related to healthcare-seeking behavior and testing practices [18]. Third, multilevel modeling accounted for clustering at the provincial level, allowing adjustment for regional variation in influenza circulation and healthcare access. The use of modified Poisson regression with robust standard errors enabled direct estimation of risk ratios, which provide a more interpretable measure of vaccine effectiveness than odds ratios when outcomes are not rare [20].

Several limitations should be acknowledged. Residual confounding may remain despite adjustment for key demographic and clinical variables. Because the analytical unit was the clinical episode and individual patients may have contributed more than one episode, within-patient correlation was not fully accounted for by provincial-level clustering, which may result in slightly underestimated standard errors for the primary VE estimates. Vaccination status was obtained from administrative records and may be subject to misclassification if vaccinations received outside the reporting system were not captured. In addition, because approximately four-fifths of study episodes occurred in men, the generalizability of these findings to women with COPD may be more limited. Information on patient-level vaccine formulation (e.g., trivalent versus quadrivalent vaccines), formal antigenic match between vaccine and circulating strains, and prior vaccination history was not available. Descriptive national information on vaccine components and circulating strains could be assembled only for a subset of study seasons and was therefore used as contextual support rather than as a formal year-by-year strain-match analysis across the full 2013–2024 study period. A complete strain-match analysis across all study seasons would have required separate reconstruction of national virologic surveillance data into a standardized season-level analytic dataset and was therefore beyond the scope of the present study. In addition, subtype-specific analyses may have limited precision in seasons with lower circulation of individual influenza strains.

### Conclusion

Overall, our findings provide national laboratory-confirmed evidence that seasonal influenza vaccination was associated with substantial protection against influenza-associated pneumonia and COPD exacerbations among patients with COPD in Thailand. These findings support continued efforts to improve influenza vaccine uptake in this high-risk population.

## Acknowledgments

The authors acknowledge the National Health Security Office and the Department of Medical Sciences, Thailand. The authors also thank Ms. Jarawee Rattanayot from the National Health Security Office for her valuable support in facilitating this project.

## DECLARATIONS

### Data Availability

The administrative healthcare data from the National Health Security Office (NHSO) used in this study are not publicly available as they contain protected health information subject to NHSO data governance policies. The laboratory-confirmed influenza surveillance data from the Department of Medical Sciences are similarly held under institutional data agreements. Researchers may submit requests for data access directly to the National Health Security Office (nhso.go.th) and the Department of Medical Sciences (dmsc.moph.go.th), subject to their respective data sharing frameworks. The analytic code used in this study is available from the corresponding author upon reasonable request.

### Author Contributions

Conceptualization, S.C., P.T., K.P., and P.P.; methodology, S.C., P.T., K.P., and P.P.; formal analysis, S.C., S.N., and P.P.; investigation, S.C. and P.T.; data curation, S.C. and S.N.; resources, P.P.; validation, P.T., J.K., K.P., and P.P.; writing—original draft preparation, S.C. and J.K.; writing—review and editing, S.C., P.T., J.K., S.N., K.P., and P.P.; visualization, J.K. and K.P.; supervision, P.P.; project administration, P.P. All authors have read and agreed to the published version of the manuscript.

## SUPPORTING INFORMATION CAPTIONS

S1 Table. Vaccine effectiveness by influenza activity level in Thailand, 2013–2024.

S2 Table. Strain-specific vaccine effectiveness among patients with chronic obstructive pulmonary disease in Thailand, 2013–2024.

S3 Table. Southern Hemisphere influenza vaccine components and yearly distribution of circulating influenza virus types in Thailand for years with available descriptive strain-context information, 2011–2015.

S4 Table. Circulating influenza strains by subtype/lineage and vaccine-strain correspondence in Thailand for years with available descriptive strain-context information, 2011–2015.

